# Urinary sodium excretion is not associated with the incidence of end-stage kidney disease and kidney-related death: results from the UK Biobank

**DOI:** 10.1101/2021.12.14.21267572

**Authors:** Ying Shan, Yong Bai, Jingwen Zhang, Yueqi Lu, Sike Yu, Congying Song, Juehan Liu, Min Jian, Junjie Xu, Zuying Xiong, Xiaoyan Huang

**Affiliations:** Clinical Research Academy, Peking University Shenzhen Hospital, Peking University, Shenzhen, China; BGI-Shenzhen, Shenzhen, China; Renal Division, Peking University Shenzhen Hospital, Peking University, Shenzhen, China

**Keywords:** cohort study, end-stage kidney disease, public health, proportional hazards models, sodium

## Abstract

**Background:** Sodium reduction lowers blood pressure and albuminuria, indicating a hypothesized but yet-to-be proven association between sodium intake and kidney-related endpoints.

**Objectives:** We aimed to investigate the associations of 24-h urinary sodium excretion, reflecting daily sodium intake, with kidney-related outcomes.

**Methods:** Prospective cohort of 444,086 middle- to early late-aged participants from the UK Biobank. The primary outcome was a composite of incident end-stage kidney disease (ESKD) and death due to a kidney-related cause, each of which was separately examined as a secondary outcome. Death due to a non-kidney related cause prior to ESKD was considered a competing event.

**Results:** The mean 24-h urinary sodium excretion estimated from spot urinary biomarkers was 3.3 g. During a median follow-up of 11.8 years, 1,256 composite events occurred. Multivariable-adjusted cause-specific hazards models showed that, with every 1-g increment in 24-h urinary sodium excretion, hazard ratios (95% confidence intervals) were 1.03 (0.91-1.16), 1.08 (0.88-1.32), and 1.01 (0.88-1.16) for the composite outcome, incident ESKD, and kidney-related death, respectively. Similar null results were observed when the exposure was treated as binary (<2 g/d vs. ≥2 g/d) or multicategorical (quartiles). Nonlinear associations were not detected with restricted cubic splines. The findings also held constant in prespecified sensitivity and subgroup analyses.

**Conclusions:** Estimated 24-h urinary sodium excretion was not linearly or nonlinearly associated with the incidence of ESKD or death due to kidney-related causes. Our findings did not support the hypothesized notion that sodium intake should be reduced to prevent kidney-related endpoints at the population level.

## INTRODUCTION

Current guidelines propose low sodium intake in adults [1], although the recommended upper limit of daily sodium intake varies between 1.5 g and 2.4 g according to different guideline-issuing authorities [2, 3]. This recommendation relies on convincing evidence that high sodium intake causally increases blood pressure, as well as on the inference that interventions to reduce blood pressure will subsequently decrease target organ damage [1]. However, whether the effects of manipulating sodium intake can successfully translate to the expected benefits remains inconclusive [4, 5]. Therefore, research that directly validates the relationship between sodium intake and hard clinical endpoints is warranted.

Uncontrolled high blood pressure has long been recognized as an important risk factor for the initiation and progression of kidney diseases. In addition, albuminuria serves as another key determinant of progressive kidney function loss [6]. Akin to its impact on blood pressure, sodium reduction ameliorates urinary excretion of albumin in people with chronic kidney disease (CKD) [7, 8]. Through causal pathways involving blood pressure and/or albuminuria, sodium intake is thus presumed to be associated with the long-term progression of kidney impairment. A recent systematic review summarizing cross-sectional, cohort, and intervention studies however concluded that robust evidence suggesting sodium reduction delays the progression of kidney function decline is lacking [7]. Previous results of this kind have been mainly obtained in patients with CKD, and notably, two major concerns exist, both of which could indicate flaws in the available evidence. Reverse causality, introduced by the fact that overt CKD patients may either eat less or adhere to the recommendation of low sodium intake, tends to lead to underestimation of the sodium-kidney outcome relation [9]. Furthermore, the ability to excrete sodium decreases with progressive decline in kidney function, compromising the reliability of urinary biomarker measurement, which is often utilized as a metric of sodium intake in epidemiological research [10]. Data from a large-scale population-based cohort would be of great value to elucidate the controversy about the hypothesized but unproven benefit of sodium reduction.

As the association between sodium intake and clinically significant kidney outcomes is still debated, we investigated the linear and nonlinear associations of 24-h urinary sodium excretion, mirroring daily sodium intake, with the incidence of end-stage kidney disease (ESKD) in 444,086 community-dwelling UK Biobank participants. In exploratory analyses, we evaluated whether these associations were mediated by blood pressure and albuminuria. Several *a priori* determined sensitivity analyses were also undertaken to confirm the main finding.

## MATERIALS AND METHODS

### Data sharing

Datasets related to this work are available at the UK Biobank resource (https://www.ukbiobank.ac.uk/), and the code used for all analyses is available on request.

### Study design and participants

The UK Biobank comprises 502,638 volunteers aged 40 to 69 years (94% of self-reported European ancestry) recruited through the United Kingdom National Health Service registers between April 2007 and December 2010. The UK Biobank received ethical approval from the North West Multi-Center Research Ethics Committee (REC reference: 11/NW/03820). Participants attended 1 of 22 dedicated assessment centers in England, Scotland, and Wales, as described elsewhere [11]. All participants gave written informed consent before enrolment, and the study was conducted in accord with the principles of the Declaration of Helsinki. After providing informed consent, participants completed a computer-based questionnaire on demographics, life-course exposures, self-reported health behavior, medical history, and treatments. Additionally, participants underwent a standardized portfolio of clinical examinations and assays of biological samples.

The present study is a prospective cohort study based on UK Biobank participants. The study hypothesis arose before inspection of the data and a research plan had been submitted to the UK Biobank (application ID 65814). However, no protocol for the present analysis was published.

Figure 1 depicts the process of inclusion and exclusion. In brief, this study included participants without ESKD (field 42026) at baseline according to a prespecified algorithm (https://biobank.ndph.ox.ac.uk/ukb/ukb/docs/alg_outcome_esrd.pdf), with random urine samples collected and tested for urine sodium and creatinine concentrations, and who did not withdraw from the UK Biobank as of August 9^th^, 2021. The exclusion criteria were incomplete data (age, sex, height, weight, etc.) for estimating 24-h urinary sodium excretion, 24-h urinary sodium excretion outliers, and prevalent malignant tumor (fields 40005 and 40012) at baseline.

**Figure 1.**
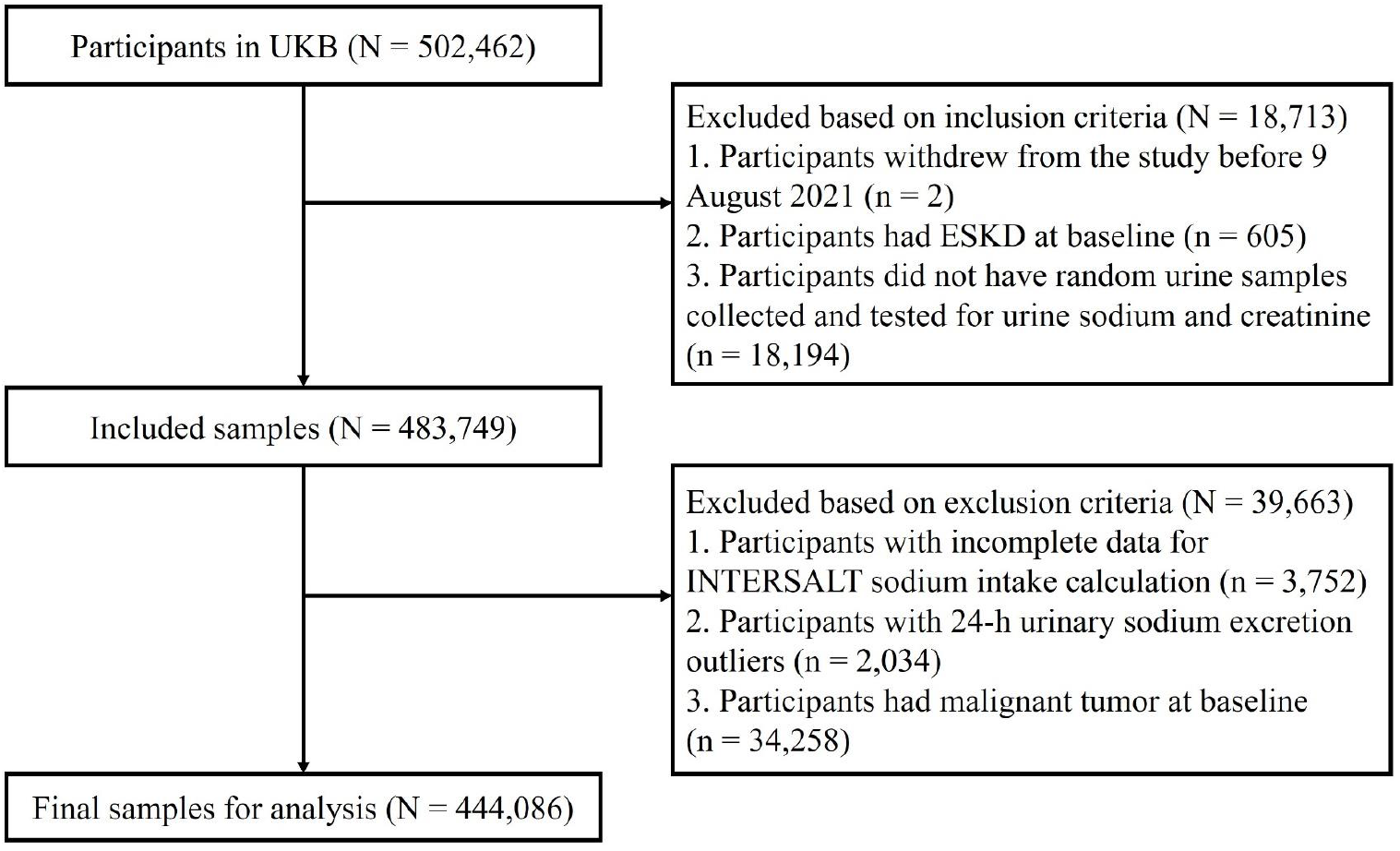
Flow chart of inclusion and exclusion. Abbreviations: ESKD, end-stage kidney disease; UKB, UK Biobank.

### Exposure estimates

A spot midstream urine sample was obtained at the end of a 2-h visit and refrigerated between 2°C and 8°C. All urinary biomarkers were measured on a single Beckman Coulter AU5400 clinical chemistry analyzer using the manufacturer’s reagents and calibrators, except for urinary albumin, for which reagents and calibrators were sourced from Randox Bioscience. The Beckman Coulter AU5400 analyzer used a potentiometric measurement for the determination of sodium (field 30530) and potassium (field 30520) concentrations and a photometric measurement for the determination of creatinine (field 30510) and albumin (field 30500) concentrations. The analysis method for urinary sodium and potassium involved a sample predilution step, while for urinary albumin and creatinine assays, it allowed samples with results exceeding the upper analytical limit of the assay to be diluted and reanalyzed. Internal quality control was performed for all 4 urinary biomarkers (http://biobank.ctsu.ox.ac.uk/crystal/docs/urine_assay.pdf).

The primary exposure was 24-h urinary sodium excretion (g/d) estimated from the spot urinary biomarker concentrations based on the sex-specific International Cooperative Study on Salt, Other Factors, and Blood Pressure (INTERSALT) equations with a Western Europe intercept [12]. The method of using spot urine samples to approximate daily excretion is widely used, especially for surveys with large populations. The INTERSALT equations, adopted in another study based on the UK Biobank data [13], are the least biased compared with other predictive equations, including the Kawasaki equations. Unlike the Kawasaki equations, the INTERSALT equations have been developed using nonfasting spot urine samples as obtained in the UK Biobank. The sex-specific Kawasaki equations were also used to estimate the exposure of interest and were used instead of the INTERSALT equations in a sensitivity analysis [14].

### Primary and secondary outcomes

The follow-up period started on the date of first assessment and ended on the date of death, first date of ESKD diagnosis, or the end of follow-up, whichever occurred first. Both retrospective and prospective data linked to electronic health records, including hospital episode statistics data on diagnoses and operations and cause of death data through the Office for National Statistics, are available in the UK Biobank. The follow-up periods ended on February 28^th^, 2021 for death data and censoring (https://biobank.ctsu.ox.ac.uk/crystal/exinfo.cgi?src=Data_providers_and_dates, last accessed October 1^st^, 2021) and on March 31^st^, 2019 for algorithmically-defined ESKD incidence (https://biobank.ctsu.ox.ac.uk/crystal/label.cgi?id=42, last accessed October 1^st^, 2021).

The primary outcome of interest was a composite endpoint consisting of ESKD and death due to a kidney-related cause. ESKD was defined as reaching CKD stage 5 or requiring kidney replacement therapy (dialysis or kidney transplantation), according to the *10*^*th*^ *revision of the International Classification of Diseases* (ICD-10) codes (E85.3, N16.5, N18.0, N18.5, Q60.1, T82.4, T86.1, Y60.2, Y61.2, Y62.2, Y84.1, Z49.0, Z49.1, Z49.2, Z94.0, and Z99.2) and the *4*^*th*^ *version of the Office of Population Censuses and Surveys Classification of Interventions and Procedures* codes (L74.1, L74.2, L74.3, L74.4, L74.5, L74.6, L74.8, L74.9, M01.2, M01.3, M01.4, M01.5, M01.8, M01.9, M02.3, M08.4, M17.2, M17.4, M17.8, M17.9, X40.1, X40.2, X40.3, X40.4, X40.5, X40.6, X40.7, X40.8, X40.9, X41.1, X41.2, X41.8, X41.9, X42.1, X42.8, X42.9, and X43.1) during hospitalization or ICD-10 codes (N18.0 and N18.5) listed in any position in a death record, as per an algorithm (https://biobank.ndph.ox.ac.uk/ukb/ukb/docs/alg_outcome_esrd.pdf) prespecified by the UK Biobank Outcome Adjudication Group. Death due to a kidney-related cause was defined as death where chronic or unspecified kidney failure was listed as a primary or contributing cause of death based on ICD-10 codes (N18 and N19) [15]. Death due to the remaining causes prior to ESKD was treated as a competing event. Additionally, we separately examined initiation of ESKD and death due to a kidney-related cause during follow-up as the secondary outcomes.

### Covariates

Covariates were 1). Demographic factors, including age (field 21022), sex (field 31), ethnicity (field 21000; categorized into white, Asian, black, and others), education (field 6138; categorized into level 1, level 2, level 3, and level 4), and Townsend deprivation index (field 189); 2). Lifestyle factors, including smoking (field 20116), alcohol consumption (field 20117), metabolic equivalents (field 22040), and 24-h urinary potassium excretion (representing daily potassium intake; calculated by the Kawasaki equation based on fields 30520 and 30510) [14]; 3). Anthropometric and clinical measurements, including body mass index (BMI; field 21001), waist circumference (field 48), systolic blood pressure (SBP; field 4080), and diastolic blood pressure (DBP; field 94); 4). Comorbid conditions, including hypertension (field 131286), diabetes (field 2443), cardiovascular diseases (CVDs; defined as any diseases of the circulatory system, excluding essential hypertension, stroke, and diseases of arteries, veins, and lymph), and stroke (field 42006); 5). Medications (field 20003), including diuretics, angiotensin-converting enzyme inhibitors (ACEIs), angiotensin II inhibitors (ARBs), and systemic corticosteroids; and 6). Kidney measures, including estimated glomerular filtration rate (eGFR; calculated by the cystatin C-based Chronic Kidney Disease Epidemiology Collaboration Equations) [16] and urine albumin-to-creatinine ratio (UACR; categorized into normal, microalbuminuria, and macroalbuminuria). Details of these measurements can be found in the UK Biobank Data Showcase (http://biobank.ctsu.ox.ac.uk/crystal/).

### Statistical analyses

We first carried out descriptive analyses. Baseline characteristics are presented as the mean (standard deviation) for symmetrical continuous variables, median (interquartile range) for skewed continuous variables, and number (percentage) for categorical variables, in the study population as a whole and according to quartiles of 24-h urinary sodium excretion. For all the endpoints, medium (interquartile range) follow-up periods and the numbers of events that occurred during the follow-up were recorded; in addition, annual incidence rates were calculated. We also used cumulative incidence function curves to describe the outcomes during follow-up, according to quartiles of 24-h urinary sodium excretion. Considering the competing risks, we plotted the cumulative incidence function curves based on both cause-specific hazards models and subdistribution hazards models.

The associations between 24-h urinary sodium excretion and outcomes were analysed. Taking into account the competing risks, we investigated the associations using cause-specific hazards models [17]. The participants lost to follow-up were considered censoring. The proportional hazard assumption was evaluated using Schoenfeld residuals. When the assumption was not fulfilled, a time-dependent covariate was constructed for continuous variables, and a stratified Cox procedure was carried out for discrete variables. Multicollinearity was assessed in each model with the variance inflation factor (VIF). The variables with VIF values above 5 were considered to have severe collinearity and were excluded except for one variable that was the most relevant to the outcomes in a biological sense.

For each of the three outcomes, we constructed a crude model and three multivariable models that included an increasing number of covariates. Model 1 was adjusted for baseline demographic (age, sex, ethnicity, education, and Townsend deprivation index) and lifestyle factors (smoking, alcohol consumption, metabolic equivalents, and 24-h urinary potassium excretion). Model 2 was adjusted for additional variables, including anthropometric measurements (BMI and waist circumference), comorbid conditions (hypertension, diabetes, CVDs, and stroke), medications (diuretics, ACEIs, ARBs, and systemic corticosteroids), and baseline eGFR. In model 3, we additionally adjusted for mediators of SBP, DBP, and UACR. The exposure of 24-h urinary sodium excretion was entered into the model as 1) a continuous variable without transformation; 2) a binary variable (<2 g/d vs. ≥2 g/d, where sodium intake of less than 2 g/d is recommended by the World Health Organization) [1]; and 3) a discrete variable constructed based on quartiles. We confirmed all these models with the Fine and Gray approach (subdistribution hazards models) as a substitute to manage the competing risks. Additionally, the potential nonlinear relationship between 24-h urinary sodium excretion and all endpoints was evaluated on a continuous scale with restricted cubic spline curves based on cause-specific hazards models. The participants with missing covariate data were excluded from the analyses. Data are presented as hazard ratios (HRs) and 95% confidence intervals (CIs).

Furthermore, to assess the robustness of the results, we performed several sensitivity analyses. First, since certain medications might interfere with the results of urinary biomarkers, we excluded participants who took diuretics, ACEIs, ARBs, or glucocorticoids at baseline. Second, to reduce the risk of reverse causation, we excluded participants who had prevalent congestive heart failure or CKD stage 4-5 at baseline, developed ESKD or died within 2 years of follow-up. Moreover, we conducted analyses using the exposure of 24-h urinary sodium excretion calculated by Kawasaki equations.

For the composite endpoint of ESKD and kidney-related death, we conducted preplanned stratified analyses to assess the potential interaction of 24-h urinary sodium excretion as a continuous variable and the following factors that might be effect modifiers or mediators: age (<50 y, 50-<60 y, and ≥60 y), sex (women or men), ethnicity (white or nonwhite), 24-h urinary potassium excretion (tertiles), hypertension (present or absent), CVDs (present or absent), stroke (present or absent), ACEIs or ARBs usage (yes or no), eGFR (<60 ml/min/1.73m^2^, 60-<90 ml/min/1.73m^2^, and ≥90 ml/min/1.73m^2^), and UACR (<30 mg/g, 30-<300 mg/g, and ≥300 mg/g). Covariates included in these analyses were in line with those in the aforementioned model 2, when appropriate. We evaluated the potential interaction by modelling the cross-product term of the stratified variable and 24-h urinary sodium excretion.

All statistical analyses were performed using R Statistical Software, version 4.0.3. All statistical tests were two-sided, and we considered a P value less than 0.05 to be statistically significant.

## RESULTS

### Baseline characteristics

Of the 502,462 UK Biobank participants, 58,376 were excluded based on the inclusion and exclusion criteria, leaving 444,086 participants eligible for analysis in the study (**Figure 1**). The mean (standard deviation) urinary sodium excretion was 3.3 (0.8) g/d calculated by INTERSALT equations and 4.1 (1.2) g/d calculated by Kawasaki equations (**Figure S1**). Only 2.1% and 2.8% of the participants met the World Health Organization recommendation that sodium intake should be less than 2 g/d, estimated by the INTERSALT and Kawasaki equations, respectively.

**Table 1** shows the baseline characteristics of the study participants collectively, as well as grouped by quartiles of 24-h urinary sodium excretion. Among the 444,086 participants, the mean age was 56.2 years, and 54.1% were women. The overall mean 24-h urinary sodium excretion was higher in men than in women (4.0 g/d vs. 2.8 g/d). In summary, people in the lowest 24-h urinary sodium excretion quartile (Q1) had a higher prevalence of white individuals and a higher education level than those in the highest quartile (Q4). In addition, people in Q1 had a higher prevalence of never smoking and never consuming alcohol. They had a lower prevalence of use of diuretics, ACEIs, or ARBs and lower rates of comorbidities, including hypertension, diabetes, CVDs, and stroke. They also had lower 24-h urinary potassium excretion, BMI, waist circumference, and blood pressure, and a lower prevalence of albuminuria.

**Table 1.** Baseline characteristics of participants.

|  | Missing<br>value, n | Whole<br>population | 24-h urinary sodium excretion |  |  |  |
| --- | --- | --- | --- | --- | --- | --- |
|  |  |  | Quartile 1 | Quartile 2 | Quartile 3 | Quartile 4 |
| Range, g/d |  |  | [0.81, 2.68] | (2.68, 3.20] | (3.20, 3.92] | (3.92, 5.84] |
| N |  | 444,086 | 111,022 | 111,021 | 111,021 | 111,022 |
| Age, year | 0 | 57 [50, 63] | 61 [54, 65] | 55 [48, 61] | 55 [48, 62] | 58 [50, 63] |
| Women, n (%) | 0 | 240,079 (54) | 106,242 (96) | 96,310 (87) | 33,200 (30) | 4,327 (4) |
| Ethnicity, n (%) | 2,077 |  |  |  |  |  |
| White |  | 417,730 (94.5) | 106,808 (96.5) | 104,200 (94.2) | 103,170 (93.4) | 103,552 (93.8) |
| Asian |  | 10,277 (2.3) | 1,440 (1.3) | 2,665 (2.4) | 3,146 (2.9) | 3,026 (2.7) |
| Black |  | 7,254 (1.7) | 1,073 (1.0) | 1,816 (1.7) | 2,293 (2.1) | 2,072 (1.9) |
| Others |  | 6,748 (1.5) | 1,301 (1.2) | 1,904 (1.7) | 1,814 (1.6) | 1,729 (1.6) |
| Townsend deprivation index | 544 | -2.14 [-3.65, 0.53] | -2.37 [-3.75, 0.01] | -2.12 [-3.63, 0.50] | -2.07 [-3.62, 0.73] | -1.96 [-3.55, 0.87] |
| Education*, n (%) | 6,469 |  |  |  |  |  |
| Level 1 |  | 73,670 (17) | 19,763 (18) | 16,171 (15) | 15,762 (15) | 21,974 (20) |
| Level 2 |  | 74,024 (17) | 20,355 (19) | 20,490 (19) | 16,634 (15) | 16,545 (15) |
| Level 3 |  | 145,253 (33) | 32,711 (30) | 35,771 (32) | 37,249 (34) | 39,522 (36) |
| Level 4 |  | 144,670 (33) | 36,724 (33) | 37,110 (34) | 39,802 (36) | 31,034 (29) |
| Metabolic equivalents,<br>minutes/week | 85,465 | 1,777 [813, 3,570] | 1,886 [916, 3,612] | 1,695 [792, 3,333] | 1,750 [792, 3,519] | 1,784 [747, 3,840] |
| Smoking status, n (%) | 2,158 |  |  |  |  |  |
| Never |  | 24,3513 (55) | 65,890 (60) | 65,268 (59) | 60,716 (55) | 51,639 (47) |
| Previous |  | 151,543 (34) | 35,267 (32) | 34,247 (31) | 36,582 (33) | 45,447 (41) |
| Current |  | 46,872 (11) | 9,389 (8) | 11,011 (10) | 13,200 (12) | 13,272 (12) |
| Alcohol consumption, n (%) | 939 |  |  |  |  |  |
| Never |  | 35,016 (8) | 9,602 (9) | 9,267 (8) | 8,523 (8) | 7,624 (7) |
| Occasional |  | 100,023 (23) | 26,836 (24) | 29,561 (27) | 23,387 (21) | 20,239 (18) |
| Frequent |  | 308,108 (69) | 74,404 (67) | 71,985 (65) | 78,846 (71) | 82,873 (75) |
| 24-h urinary potassium excretion, g/d | 0 | 2.75 ± 0.59 | 2.60 ± 0.57 | 2.67 ± 0.56 | 2.81 ± 0.60 | 2.93 ± 0.58 |
| Body mass index, kg/m<sup>2</sup> | 0 | 27.4 ± 4.7 | 24.6 ± 3.6 | 27.1 ± 4.1 | 27.8 ± 4.9 | 30.0 ± 4.5 |
| Waist circumference, cm | 76 | 90 ± 13 | 80 ± 10 | 86 ± 11 | 93 ± 10 | 101 ± 11 |
| Hypertension, n (%) | 2 | 42,545 (10) | 8,486 (8) | 8,626 (8) | 10,036 (9) | 15,397 (14) |
| Diabetes, n (%) | 1,868 | 22,336 (5) | 2,835 (3) | 3,892 (4) | 5,708 (5) | 9,901 (9) |
| Cardiovascular disease, n (%) | 0 | 39,614 (9) | 7,981 (7) | 7,046 (6) | 10,292 (9) | 14,295 (13) |
| Stroke, n (%) | 127 | 6,314 (1.4) | 1,397 (1.3) | 1,203 (1.1) | 1,574 (1.4) | 2,140 (2.0) |
| Diuretics, n (%) | 0 | 31,779 (7) | 6,463 (6) | 7,692 (7) | 6,906 (6) | 10,718 (10) |
| ACEIs or ARBs, n (%) | 0 | 69,143 (16) | 12,366 (11) | 12,926 (12) | 16,687 (15) | 27,164 (25) |
| Systolic blood pressure, mmHg | 24,961 | 138 ± 19 | 135 ± 19 | 135 ± 19 | 138 ± 17 | 143 ± 17 |
| Diastolic blood pressure, mmHg | 24,958 | 82 ± 10 | 79 ± 10 | 81 ± 10 | 83 ± 10 | 86 ± 10 |
| eGFR, ml/min/1.73m<sup>2</sup> | 25,313 | 88.8 ± 16 | 87.5 ± 15.3 | 90.8 ± 15.8 | 89.9 ± 16.0 | 87.1 ± 16.3 |
| UACR, n (%) | 14,049 |  |  |  |  |  |
| <30 mg/g |  | 410,572 (95.5) | 100,610 (95.9) | 101,036 (96.0) | 104,972 (95.8) | 103,954 (94.2) |
| 30-< 300mg/g |  | 17,821 (4.1) | 4,048 (3.9) | 3,898 (3.7) | 4,121 (3.8) | 5,754 (5.2) |
| ≥300 mg/g |  | 1,644 (0.4) | 277 (0.2) | 300 (0.3) | 412 (0.4) | 655 (0.6) |

Mean ± standard deviation, median [interquartile range], and frequency (percentage) are shown for symmetrical continuous variables, skewed continuous variables, and categorical variables, respectively.
\*Level 1, “none of the above”; level 2, “O levels/GCSEs or equivalent” or “CSEs or equivalent”; level 3, “A levels/AS levels or equivalent” or “NVQ or HND or HNC or equivalent” or “Other professional qualifications”; level 4, “College or University degree”.
Abbreviations: ACEIs, angiotensin-converting enzyme inhibitors; ARBs, angiotensin II receptor blockers; eGFR, estimated glomerular filtration rate; UACR, urine albumin-to-creatinine ratio.

### 24-h urinary sodium excretion and the outcomes

The mean follow-up period for the composite endpoint of ESKD and kidney-related death was 11.8 (interquartile range 11.3-12.7) years, that for incident ESKD was 10.0 (interquartile range 9.4-10.8) years, and that for kidney mortality was 11.8 (interquartile range 11.3-12.7) years. During follow-up, 1130 (0.3%) participants lost to follow-up, 1,256 (0.3%) participants developed the composite endpoint, 459 (0.1%) participants progressed to ESKD, and 962 (0.2%) participants died due to a kidney-related cause. The incidence rates were 24.0, 10.3, and 18.3 per 100,000 person-years for the composite endpoint, ESKD, and kidney-related death, respectively.

**Figure 2** presents the cumulative incidence functions for participants in quartiles of 24-h urinary sodium excretion based on cause-specific and subdistribution hazards models. Both approaches provided similar estimated cumulative incidence functions for all the outcomes. By 12.7 (Q3 of the follow-up time) years, cumulative incidence rates of primary and secondary outcomes were higher in participants in Q4 (0.6%) than those in Q1 (0.2%).

**Figure 2.**
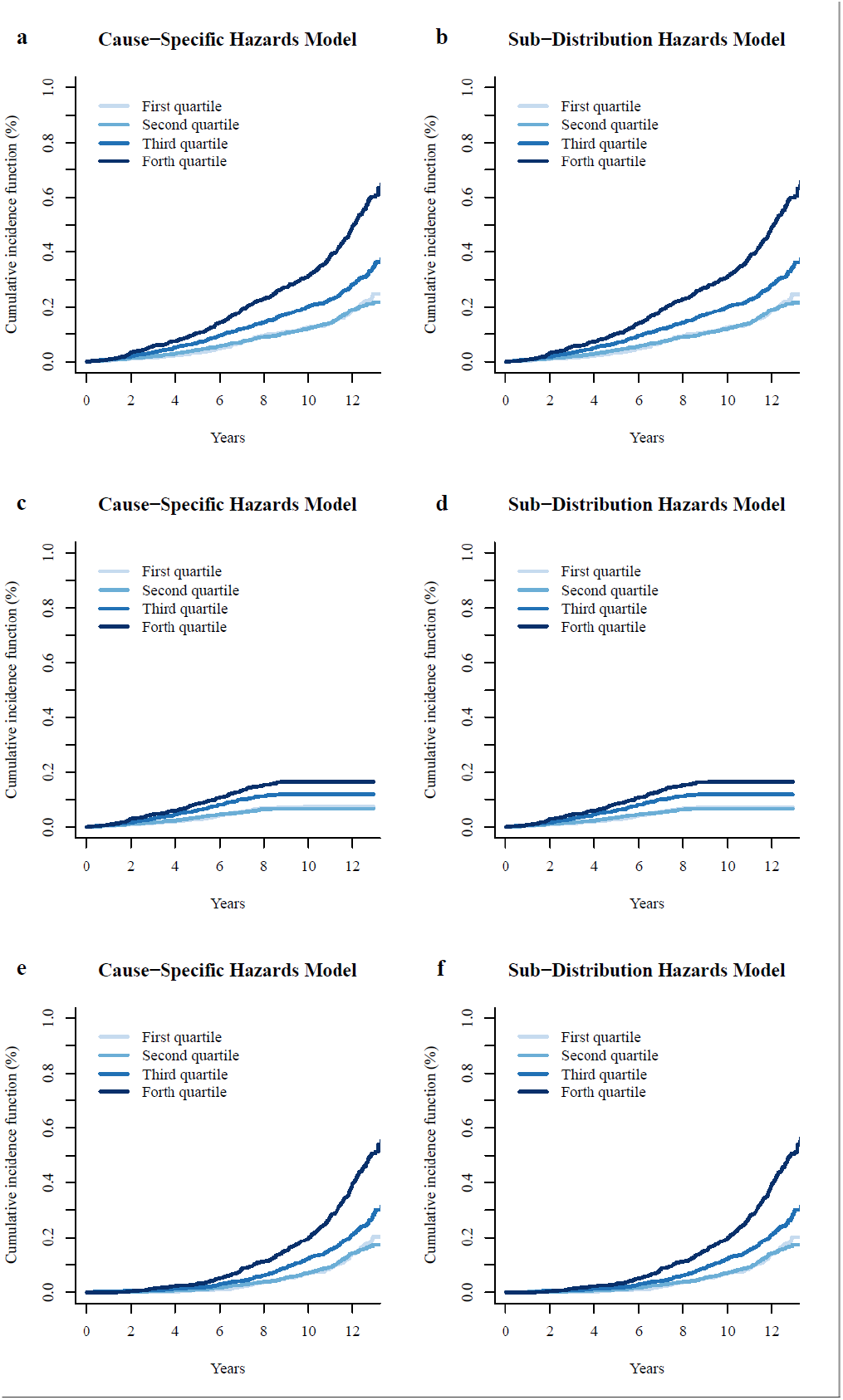
Cumulative incidence functions for primary (a and b) and secondary outcomes (c and d for end-stage kidney disease; e and f for kidney-related death) by 24-h sodium excretion quartiles on the basis of cause-specific hazards models and subdistribution hazards models.

As shown in **Table 2**, in crude models, 24-h urinary sodium excretion appeared to be positively associated with the composite endpoint of ESKD and kidney-related death and with each component. Next, we examined the association between 24-h urinary sodium excretion and the primary and secondary outcomes, adjusted for potential confounders, including age, sex, ethnicity, education, Townsend deprivation index, smoking, alcohol consumption, metabolic equivalents, and 24-h urinary potassium excretion in model 1, and for waist circumference, hypertension, diabetes, CVDs, stroke, diuretics, ACEIs, ARBs, systemic corticosteroids, and eGFR in model 2 (BMI was excluded due to the collinearity problem). The cause-specific hazards results shown in **Table 2** indicate that associations between all forms of 24-h urinary sodium excretion and the composite endpoint were weak and statistically nonsignificant. Likewise, null results were observed for the secondary outcomes separately. The visualized results shown in **Figure 3** are consistent with these analyses. The HRs for the outcomes were not significantly different from one, with the confidence interval overlapping the null across the spectrum of 24-h urinary sodium excretion. After additionally adjusting for the mediators of SBP, DBP, and UACR in model 3, the results generally held null for the outcomes, and the HRs of 24-h urinary sodium excretion on a continuous scale further decreased somewhat compared with those in model 2 (**Table 2**). Results were also confirmed with subdistribution hazards models to address the competing risks, as shown in **Table S1**.

**Table 2.** Incidence rates and adjusted hazard ratios for primary and secondary outcomes according to 24-h urinary sodium excretion with cause-specific hazards models.

|  | Incidence rate,<br>per 100,000<br>person-years | Crude |  | Model 1 |  | Model 2 |  | Model 3 |  |
| --- | --- | --- | --- | --- | --- | --- | --- | --- | --- |
|  |  | HRs (95% CIs) | P | HRs (95% CIs) | P | HRs (95% CIs) | P | HRs (95% CIs) | P |
| ***Composite of end-stage kidney disease and kidney-related death*** |  |  |  |  |  |  |  |  |  |
| Continuous, per 1 g/d increment | 23.96 | 1.68 (1.58, 1.79) | <0.001 | 1.43 (1.28, 1.60) | <0.001 | 1.03 (0.91, 1.16) | 0.62 | 0.99 (0.86, 1.13) | 0.85 |
| Binary |  |  |  |  |  |  |  |  |  |
| Below 2 g/d | 30.91 | Reference |  | Reference |  | Reference |  | Reference |  |
| Above 2 g/d | 23.81 | 0.77 (0.55, 1.08) | 0.13 | 0.78 (0.52, 1.17) | 0.24 | 0.85 (0.56, 1.27) | 0.42 | 0.72 (0.47, 1.12) | 0.14 |
| Multicategory |  |  |  |  |  |  |  |  |  |
| Quartile 1 | 15.63 | Reference |  | Reference |  | Reference |  | Reference |  |
| Quartile 2 | 15.23 | 0.97 (0.80, 1.18) | 0.74 | 1.31 (1.03, 1.68) | 0.03 | 1.00 (0.78, 1.29) | 0.98 | 1.05 (0.80, 1.39) | 0.71 |
| Quartile 3 | 23.74 | 1.52 (1.27, 1.81) | <0.001 | 1.38 (1.04, 1.82) | 0.03 | 1.05 (0.80, 1.38) | 0.74 | 1.03 (0.76, 1.40) | 0.83 |
| Quartile 4 | 41.43 | 2.64 (2.25, 3.10) | <0.001 | 1.86 (1.39, 2.50) | <0.001 | 1.08 (0.80, 1.47) | 0.62 | 0.97 (0.69, 1.37) | 0.87 |
| P for linear trend |  | <0.001 |  | <0.001 |  | 0.60 |  | 0.95 |  |
| P for quadratic trend |  | <0.001 |  | 0.84 |  | 0.85 |  | 0.50 |  |
| ***End-stage kidney disease*** |  |  |  |  |  |  |  |  |  |
| Continuous, per 1 g/d increment | 10.34 | 1.57 (1.41, 1.75) | <0.001 | 1.31 (1.09, 1.56) | 0.003 | 1.08 (0.88, 1.32) | 0.47 | 0.96 (0.76, 1.20) | 0.70 |
| Binary |  |  |  |  |  |  |  |  |  |
| Below 2 g/d | 6.43 | Reference |  | Reference |  | Reference |  | Reference |  |
| Above 2 g/d | 10.43 | 1.62 (0.72, 3.63) | 0.24 | 1.82 (0.67, 4.93) | 0.24 | 2.05 (0.75, 5.59) | 0.16 | 1.66 (0.60, 4.54) | 0.33 |
| Multicategory |  |  |  |  |  |  |  |  |  |
| Quartile 1 | 7.12 | Reference |  | Reference |  | Reference |  | Reference |  |
| Quartile 2 | 6.54 | 0.92 (0.67, 1.27) | 0.62 | 1.13 (0.77, 1.65) | 0.54 | 0.91 (0.61, 1.36) | 0.65 | 1.00 (0.65, 1.53) | 0.98 |
| Quartile 3 | 11.64 | 1.64 (1.24, 2.17) | <0.001 | 1.25 (0.81, 1.91) | 0.31 | 1.10 (0.72, 1.69) | 0.66 | 1.00 (0.62, 1.62) | 0.99 |
| Quartile 4 | 16.12 | 2.27 (1.74, 2.96) | <0.001 | 1.38 (0.88, 2.16) | 0.16 | 1.08 (0.65, 1.76) | 0.78 | 0.85 (0.49, 1.46) | 0.55 |
| P for linear trend |  | <0.001 |  | 0.17 |  | 0.63 |  | 0.60 |  |
| P for quadratic trend |  | 0.04 |  | 0.95 |  | 0.78 |  | 0.51 |  |
| ***Kidney-related death*** |  |  |  |  |  |  |  |  |  |
| Continuous, per 1 g/d increment | 18.34 | 1.76 (1.64, 1.89) | <0.001 | 1.52 (1.33, 1.72) | <0.001 | 1.01 (0.88, 1.16) | 0.91 | 0.98 (0.84, 1.14) | 0.82 |
| Binary |  |  |  |  |  |  |  |  |  |
| Below 2 g/d | 27.26 | Reference |  | Reference |  | Reference |  | Reference |  |
| Above 2 g/d | 18.15 | 0.66 (0.46, 0.95) | 0.03 | 0.66 (0.43, 1.00) | 0.05 | 0.67 (0.44, 1.03) | 0.07 | 0.60 (0.38, 0.95) | 0.03 |
| Multicategory |  |  |  |  |  |  |  |  |  |
| Quartile 1 | 11.51 | Reference |  | Reference |  | Reference |  | Reference |  |
| Quartile 2 | 11.21 | 0.96 (0.77, 1.21) | 0.74 | 1.38 (1.03, 1.84) | 0.03 | 1.01 (0.75, 1.37) | 0.92 | 1.13 (0.81, 1.56) | 0.47 |
| Quartile 3 | 17.93 | 1.55 (1.27, 1.91) | <0.001 | 1.56 (1.12, 2.18) | 0.009 | 1.07 (0.78, 1.48) | 0.66 | 1.18 (0.82, 1.68) | 0.37 |
| Quartile 4 | 32.87 | 2.85 (2.36, 3.43) | <0.001 | 2.17 (1.54, 3.07) | <0.001 | 1.05 (0.73, 1.50) | 0.80 | 1.04 (0.70, 1.54) | 0.85 |
| P for linear trend |  | <0.001 |  | <0.001 |  | 0.73 |  | 0.81 |  |
| P for quadratic trend |  | <0.001 |  | 0.94 |  | 0.81 |  | 0.20 |  |

Model 1 was adjusted for age, sex, Townsend deprivation index, education, ethnicity, smoking status, alcohol consumption, metabolic equivalents, and 24-h urinary potassium excretion.
Model 3 was adjusted for age, sex, Townsend deprivation index, education, ethnicity, smoking status, alcohol consumption, metabolic equivalents, 24-h urinary potassium excretion, waist circumference, hypertension, diabetes, cardiovascular disease, stroke, diuretics, angiotensin-converting enzyme inhibitors or angiotensin II receptor blockers, estimated glomerular filtration rate, systolic blood pressure, diastolic blood pressure, and urine albumin-to-creatinine ratio.
Abbreviations: CIs, confidence intervals; HRs, hazard ratios.

**Figure 3.**
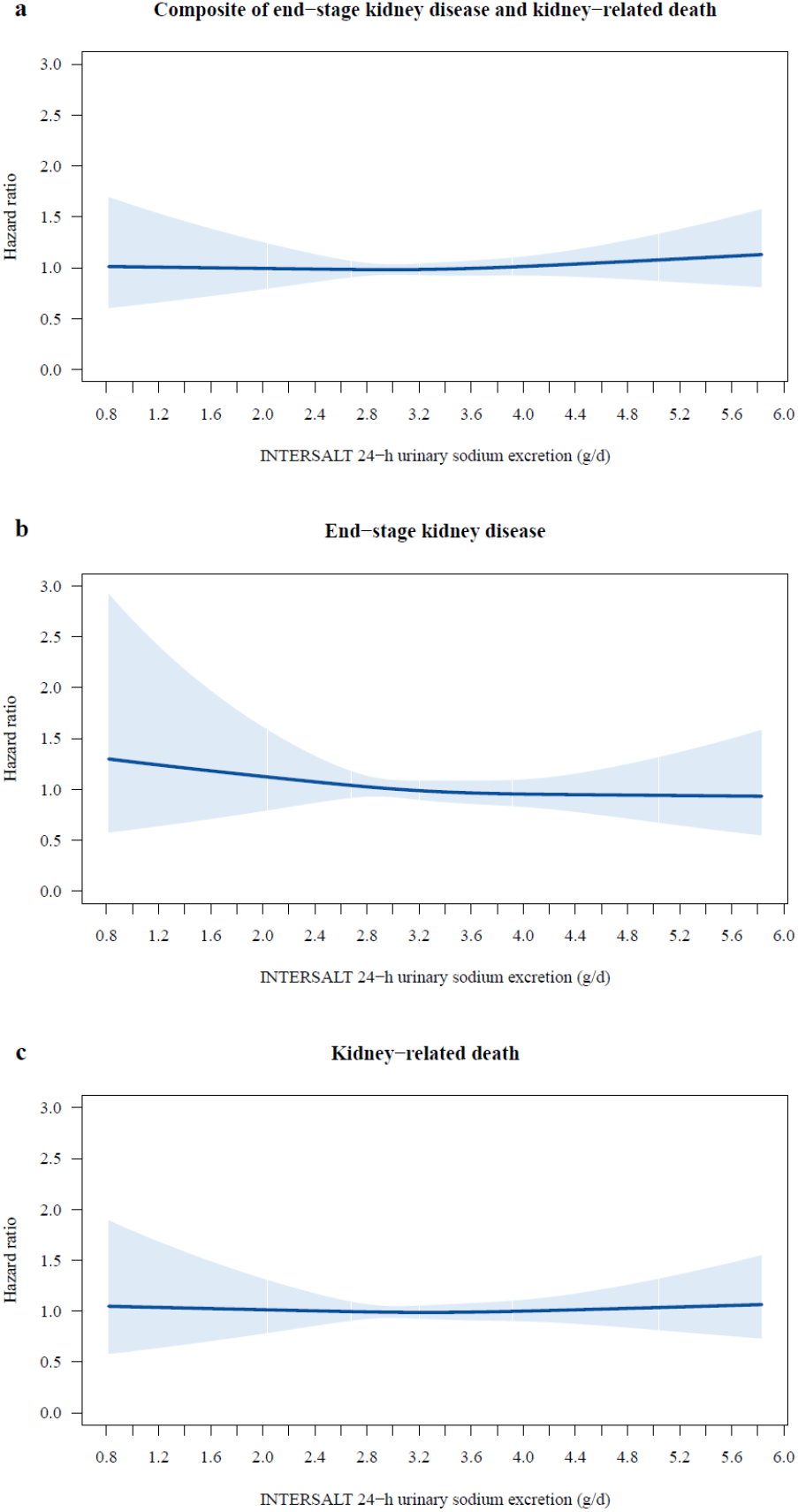
Restricted cubic splines for primary (a) and secondary outcomes (b for end-stage kidney disease, and c for kidney caused death). The analyses were adjusted for age, sex, ethnicity, education, Townsend deprivation index, smoking, alcohol consumption, metabolic equivalents, 24-h urinary potassium excretion, waist circumference, hypertension, diabetes, cardiovascular disease, stroke, diuretics, angiotensin-converting enzyme inhibitors, angiotensin II receptor blockers, systemic corticosteroids, and estimated glomerular filtration rate. Each point of the curve is the pointwise average hazard ratio. Shaded areas represent 95% confidence intervals. White vertical lines through confidence shade represent 2.5^th^ percentile, first quartile, median, third quartile, and 97.5^th^ percentile of 24-h urinary sodium excretion.

### Sensitivity and subgroup analyses

The result that 24-h urinary sodium excretion was not significantly associated with the composite endpoint of ESKD and renal caused death was also found in the following sensitivity analyses: 1) when we excluded participants who used diuretics, ACEIs, ARBs, or glucocorticoids at baseline; 2) when we excluded participants who had congestive heart failure or CKD stage 4-5 at baseline or developed ESKD or died within two years of follow-up; and 3) when we used the exposure of 24-h urinary sodium excretion calculated by the Kawasaki equations (**Table S2**).

We conducted stratified analyses for the primary outcome of interest according to several prespecified factors. We observed that in all the subgroups, the HRs of 24-h urinary sodium excretion were not significantly different from one (**Figure 4**). In addition, the P > 0.05 of all cross-product terms did not support that the effect size of 24-h urinary sodium excretion on the composite endpoint was significantly different in each stratum.

**Figure 4.**
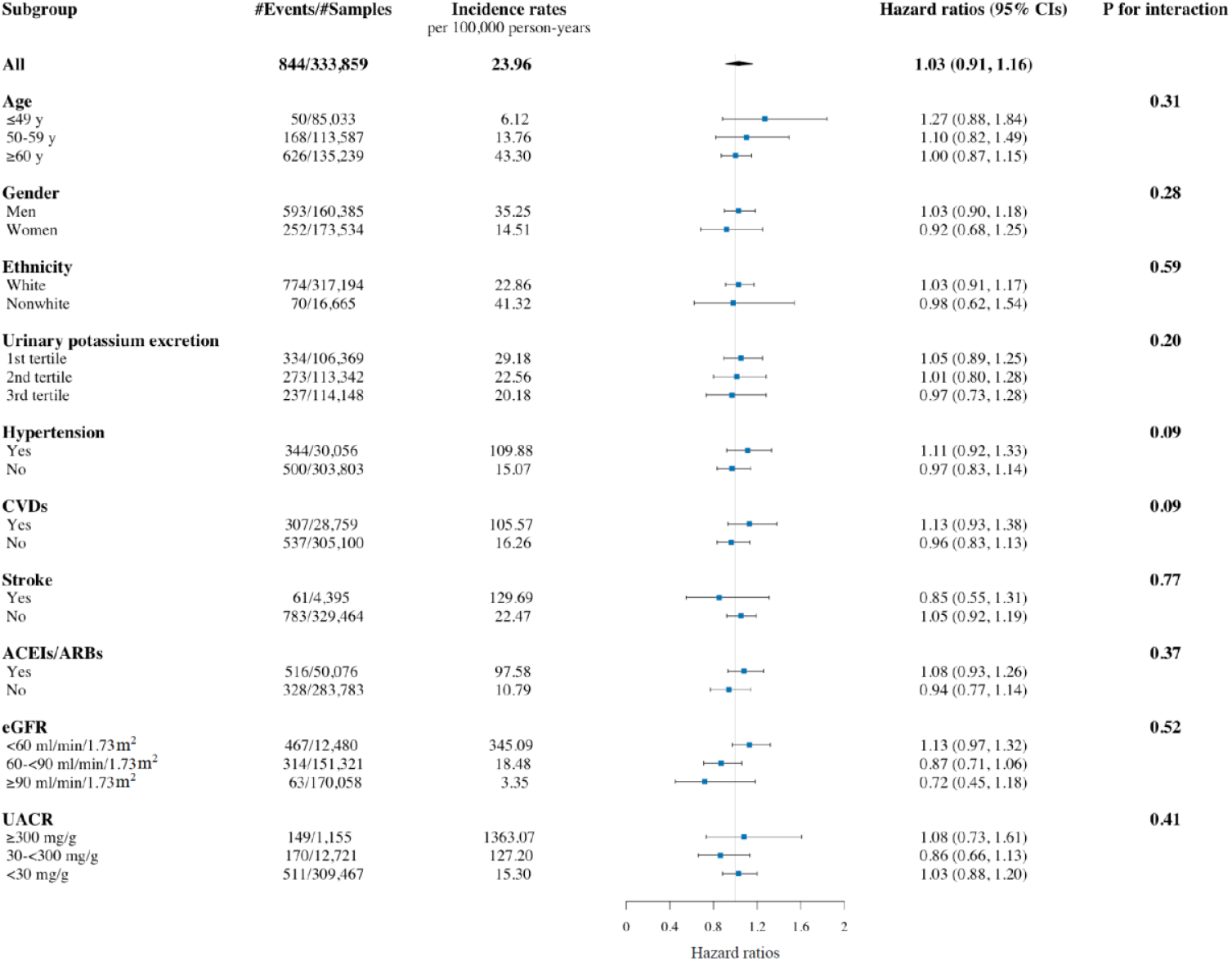
Hazard ratio for the primary outcome in prespecified subgroups. The analyses were adjusted for age, sex, Townsend deprivation index, education, ethnicity, smoking status, alcohol consumption, metabolic equivalents, 24-h urinary potassium excretion, waist circumference, hypertension, diabetes, cardiovascular disease, stroke, diuretics, angiotensin-converting enzyme inhibitors or angiotensin II receptor blockers, and estimated glomerular filtration rate, if appropriate. Abbreviations: ACEIs, angiotensin-converting enzyme inhibitors; ARBs, angiotensin II receptor blockers; CIs, confidence intervals; CVDs, cardiovascular disease; eGFR, estimated glomerular filtration rate; UACR, urine albumin-to-creatinine ratio.

## DISCUSSION

In this large prospective population-based cohort study, we observed no associations, linear or nonlinear, between estimated 24-h urinary sodium excretion and future kidney-related hard endpoints in 444,086 UK Biobank participants followed for over 10 years. This study is the first to investigate the association between sodium intake and kidney-related endpoints in the general population. The solid null results add to the literature that sodium restriction does not suffice to translate to clinically significant reductions in kidney-related events.

A number of strengths contribute to the significance of the present work. This study utilized the largest scale, prospective data to date to investigate the associations of urinary sodium excretion with ESKD and kidney-related death. The relatively large number of events witnessed during follow-up provided us with the possibility of addressing this issue with sufficient power, assuring the validity of our negative results. The nature of the population-based cohort, combining with further exclusion of participants with overt or possibly subclinical morbid conditions that readily altered sodium intake at baseline in a sensitivity analysis, reduced the risk of reverse causality. Strict quality control for data (and biological samples) collection and outcome adjudication minimized systematic errors. Last, the use of both linear and nonlinear models, the consideration of competing risks, and a series of sensitivity analyses improve the robustness and consistency of the main finding.

Overall, randomized trials have shown that sodium restriction (< 2.3 g/d) reduces blood pressure and albuminuria in CKD and general populations [18-21], without a beneficial effect on kidney function being reported [19-24]. Nevertheless, these interventional studies are typically limited by sample size and short duration. A recent Cochrane systematic review evaluated 21 randomized controlled trials published through October 2020, involving a total of 1197 adults with CKD (725 in low sodium vs. 725 in high sodium) [8]. The average study duration was only 7 weeks, ranging from 1 to 36 weeks. None of these trials assessed the impact on the incidence of death or the need for kidney replacement therapy. Instead, these studies focused on surrogate markers, for instance, blood pressure, proteinuria, serum creatinine, or body weight. More recently, the results from a large cluster-randomized trial including 20,995 participants at high risk of stroke did not support a favorable effect of salt substitutes on kidney-related death, which was treated as one of the secondary endpoints, during 5 years of follow-up [25]. As a result, intervention studies to date have provided little evidence directly linking sodium restriction with long-term kidney outcomes, albeit the findings of lowering blood pressure and albuminuria in the short term are of high certainty.

The results of kidney disease progression therefore have to be largely derived from observational studies. Previous publications, however, show conflicting results investigating the association between sodium intake and subsequent onset or progression of CKD. Several studies observed that high sodium intake is associated with future ESKD, halving of eGFR from baseline, or risk of death [26-29], but the findings could not be replicated by others [30-32]. For this reason, limited and inconsistent evidence supports an association between high sodium intake and kidney outcomes [7, 33]. The large discrepancy in findings in the CKD population is attributable, at least to some extent, to reverse causality and measurement errors. Distinct from randomized trials, sodium intake, usually measured using urinary biomarkers or dietary recall, is the exposure factor in observational studies, and subjects are divided according to the levels of sodium intake rather than randomized assignment. Patients with CKD may eat less due to poor appetite or adherence to the recommendations, resulting in reverse causality [9]. With a decline in kidney function, the ability to excrete sodium is likely to be compromised, further complicating the relationship of urinary sodium-based exposure with outcomes [10]. Data in the non-CKD population are less prone to the aforementioned bias. Even though somewhat mixed [34], most results propose that high sodium intake is associated with developing CKD in high-risk individuals [35-37], as well as with longitudinal changes in eGFR [38, 39]. Nevertheless, the relatively small sample size may have prevented researchers from further determining the association between sodium intake and ESKD or kidney-related mortality in the previous literature [40, 41].

Apart from observational and interventional studies, Mendelian randomization, which utilizes genetic variants as instrumental variable [42], also contributes to causal inference within a triangulation framework [43]. In a two-sample Mendelian randomization analysis, high sodium intake was not unfavorably linked with kidney function [44]. Since each study approach has different key sources of potential bias that are unrelated to each other [45], the reality that the results of interventional and Mendelian randomization studies altogether point to a similar null conclusion [25, 44] indeed strengthens confidence in our observational findings.

We found that sodium intake is not a risk factor, or is solely a minor risk factor compared with other established risk factors for incident ESKD or death. The reasons have not been fully elucidated. One possible interpretation is, as depicted in **Figure 5**, that sodium restriction leads to a reduction in levels of blood pressure and albuminuria but comes at the cost of higher plasma renin and aldosterone concentrations. Evidence of such side effects is consistent, indicating a compensatory response to a decrease in effective circulating volume caused by sodium restriction, especially in persons with normal blood pressure [46]. Both renin [47, 48] and aldosterone [49] can directly cause kidney damage, in turn counteracting the potential benefits via favorable effects of sodium restriction on blood pressure and albuminuria. Specifically, our study cohort mostly consisted of participants at low- or intermediate-risk of future kidney-related events, with approximately 90% and 95% of them being normotensive and normoalbuminuric, respectively. The net effect of sodium intake tended to be neutral after balancing its beneficial and detrimental effects, as observed in such a community-dwelling population. Another interpretation is that only a single baseline measurement of the exposure was available in most of the participants, precluding us from implementing the time-varying covariate. It has been reported that approximately half of people might alter their dietary sodium intake by more than 0.8 g over time [50]. On the other hand, some previous studies have proposed that both high sodium and low sodium may be diversely associated with clinical outcomes, and the effect of each may negate the effect of the other. However, this suspicion is not supported by our nonlinear analysis.

**Figure 5.**
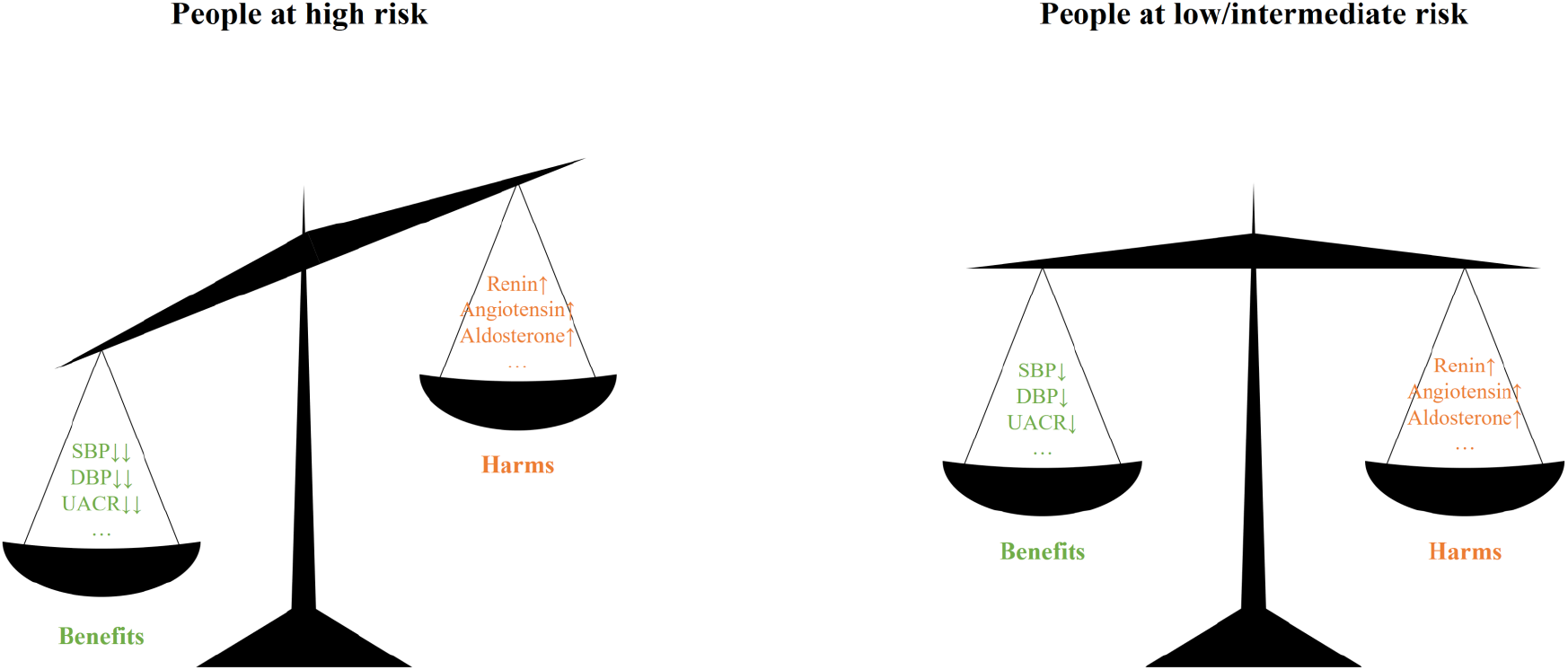
Proposed interpretation of the net effects of sodium restriction in different people. Sodium restriction leads to a reduction in levels of blood pressure and albuminuria but comes at the cost of certain adverse risks, such as higher plasma renin and aldosterone concentrations. In people at high risk, the benefits of sodium reduction exceed the harms; whereas the harms may counteract the benefits in people at low- or intermediate risk. Abbreviations: DBP, diastolic blood pressure; SBP, systolic blood pressure; UACR, urine albumin-to-creatinine ratio.

Several limitations of our study should be acknowledged. First, given the observational nature of the data, we described associations rather than inferring causality because reverse causality and residual confounding were impossible to completely avoid. Second, participants recruited to the UK Biobank were volunteers and, thus, may not be representative of the general population. Additionally, the vast majority of participants were of European white descent despite the inclusion of other ethnicities. Hence, our results should be interpreted cautiously and await confirmation in other ethnic groups with significantly different diets or prevalence of and predispositions to kidney diseases. Third, a random urinary spot sample was used as a measure of sodium excretion due to the difficulty in collecting and processing overnight or 24-h urine samples in such a very large population cohort with multiple study centers and a central biobank. We had to employ both the INTERSALT and Kawasaki equations, which led to cross-validation to some extent since the bias of both approaches may not be identical [51]. Sodium intake appeared linearly associated with blood pressure and urine albumin-to-creatinine ratio, i.e., positive controls, reducing the probability that exposure misclassification biased results to the null. Lastly, repeat eGFR measures over the follow-up period were not available for most UK Biobank participants, preventing us from further investigating the association of sodium intake with trajectories of kidney function decline.

## CONCLUSIONS

To conclude, this population-based cohort study suggested that estimated 24-h urinary sodium excretion was not linearly or nonlinearly associated with the incidence of ESKD or death due to a kidney-related cause. Compared to international recommendations and guidelines, the UK Biobank participants, in line with the world’s population in general, consume too much dietary sodium [52]. Our finding nonetheless did not support the hypothesized notion that sodium intake should be reduced to prevent the development and progression of fatal and nonfatal kidney events, at least in individuals at low- or intermediate-risk. It might be more appropriate for future guidelines to propose tailored range of sodium consumption.

## Supporting information

SUPPLEMENTAL TABLE OF CONTENTS

## Data Availability

Datasets related to this work are available at the UK Biobank resource, and the code used for all analyses is available on request.

https://www.ukbiobank.ac.uk/

## ACKNOWLEDGMENTS

This research was conducted using the UK Biobank Resource under Application Number 65814. We thank all participants of the UK Biobank. JZ, XZ, and XH were supported by a “San-ming” Project of Medicine in Shenzhen (project ID SZSM201812097). XH was supported by the Shenzhen Science and Technology Innovation Commission (grant No JCYJ20200109140412476) and Peking University Shenzhen Hospital (grant No LCYJ2020001). The funders had no role in the study design; in the collection, analysis, and interpretation of data; in the writing of the report; or in the decision to submit the article for publication.

## SUPPLEMENTAL TABLE OF CONTENTS

**Figure S1**. Histograms of 24-h urinary potassium excretion calculated by the INTERSALT (a) and Kawasaki (b) equations.

Vertical dashed lines indicate the upper limit of daily sodium intake (2 g/d) recommended by the World Health Organization.

**Table S1**. Incidence rates and adjusted hazard ratios for primary and secondary outcomes according to 24-h urinary sodium excretion with subdistribution hazards models.

**Table S2**. Adjusted hazard ratios for primary outcomes according to 24-h urinary sodium excretion under 3 sensitivity analyses.

## REFERENCES

1 World Health Organization. (2012). Guideline: sodium intake for adults and children. World Health Organization. https://apps.who.int/iris/handle/10665/77985

2 Visseren FLJ, Mach F, Smulders YM, et al. 2021 ESC Guidelines on cardiovascular disease prevention in clinical practice. Eur Heart J 2021; 42: 3227–337.

3 Whelton PK, Appel LJ, Sacco RL, et al. Sodium, blood pressure, and cardiovascular disease: further evidence supporting the American Heart Association sodium reduction recommendations. Circulation 2012; 126: 2880–9.

4 O’Donnell M, Mente A, Alderman MH, et al. Salt and cardiovascular disease: insufficient evidence to recommend low sodium intake. Eur Heart J 2020; 41: 3363–73.

5 Cook NR, He FJ, MacGregor GA, Graudal N. Sodium and health-concordance and controversy. BMJ 2020; 369: m2440.

6 Levin A, Stevens PE. Summary of KDIGO 2012 CKD Guideline: behind the scenes, need for guidance, and a framework for moving forward. Kidney Int 2014; 85: 49–61.

7 Nomura K, Asayama K, Jacobs L, Thijs L, Staessen JA. Renal function in relation to sodium intake: a quantitative review of the literature. Kidney Int 2017; 92: 67–78.

8 McMahon EJ, Campbell KL, Bauer JD, Mudge DW, Kelly JT. Altered dietary salt intake for people with chronic kidney disease. Cochrane Database Syst Rev 2021; 6: CD010070.

9 Burnier M. Sodium intake and progression of chronic kidney disease-has the time finally come to do the impossible: a prospective randomized controlled trial? Nephrol Dial Transplant 2021; 36: 381–4.

10 Shemin D, Dworkin LD. Sodium balance in renal failure. Curr Opin Nephrol Hypertens 1997; 6: 128–32.

11 Sudlow C, Gallacher J, Allen N, et al. UK biobank: an open access resource for identifying the causes of a wide range of complex diseases of middle and old age. PLoS Med 2015; 12: e1001779.

12 Brown IJ, Dyer AR, Chan Q, et al. Estimating 24-hour urinary sodium excretion from casual urinary sodium concentrations in Western populations: the INTERSALT study. Am J Epidemiol 2013; 177: 1180–92.

13 Elliott P, Muller DC, Schneider-Luftman D, et al. Estimated 24-Hour Urinary Sodium Excretion and Incident Cardiovascular Disease and Mortality Among 398 628 Individuals in UK Biobank. Hypertension 2020; 76: 683–91.

14 Kawasaki T, Itoh K, Uezono K, Sasaki H. A simple method for estimating 24 h urinary sodium and potassium excretion from second morning voiding urine specimen in adults. Clin Exp Pharmacol Physiol 1993; 20: 7–14.

15 Smyth A, Griffin M, Yusuf S, et al. Diet and Major Renal Outcomes: A Prospective Cohort Study. The NIH-AARP Diet and Health Study. J Ren Nutr 2016; 26: 288–98.

16 Inker LA, Schmid CH, Tighiouart H, et al. Estimating glomerular filtration rate from serum creatinine and cystatin C. N Engl J Med 2012; 367: 20–9.

17 Hsu JY, Roy JA, Xie D, et al. Statistical Methods for Cohort Studies of CKD: Survival Analysis in the Setting of Competing Risks. Clin J Am Soc Nephrol 2017; 12: 1181–9.

18 McMahon EJ, Bauer JD, Hawley CM, Isbel NM, Stowasser M, Johnson DW, Campbell KL. A randomized trial of dietary sodium restriction in CKD. J Am Soc Nephrol 2013; 24: 2096–103.

19 Meuleman Y, Hoekstra T, Dekker FW, et al. Sodium Restriction in Patients With CKD: A Randomized Controlled Trial of Self-management Support. Am J Kidney Dis 2017; 69: 576–86.

20 Vogt L, Waanders F, Boomsma F, de Zeeuw D, Navis G. Effects of dietary sodium and hydrochlorothiazide on the antiproteinuric efficacy of losartan. J Am Soc Nephrol 2008; 19: 999–1007.

21 Suckling RJ, He FJ, Markandu ND, MacGregor GA. Modest Salt Reduction Lowers Blood Pressure and Albumin Excretion in Impaired Glucose Tolerance and Type 2 Diabetes Mellitus: A Randomized Double-Blind Trial. Hypertension 2016; 67: 1189–95.

22 Saran R, Padilla RL, Gillespie BW, et al. A Randomized Crossover Trial of Dietary Sodium Restriction in Stage 3-4 CKD. Clin J Am Soc Nephrol 2017; 12: 399–407.

23 de Vries LV, Dobrowolski LC, van den Bosch JJ, et al. Effects of Dietary Sodium Restriction in Kidney Transplant Recipients Treated With Renin-Angiotensin-Aldosterone System Blockade: A Randomized Clinical Trial. Am J Kidney Dis 2016; 67: 936–44.

24 Bovee DM, Visser WJ, Middel I, et al. A Randomized Trial of Distal Diuretics versus Dietary Sodium Restriction for Hypertension in Chronic Kidney Disease. J Am Soc Nephrol 2020; 31: 650–62.

25 Neal B, Wu Y, Feng X, et al. Effect of Salt Substitution on Cardiovascular Events and Death. N Engl J Med 2021; 385: 1067–77.

26 Thomas MC, Moran J, Forsblom C, et al. The association between dietary sodium intake, ESRD, and all-cause mortality in patients with type 1 diabetes. Diabetes Care 2011; 34: 861–6.

27 He J, Mills KT, Appel LJ, et al. Urinary Sodium and Potassium Excretion and CKD Progression. J Am Soc Nephrol 2016; 27: 1202–12.

28 Vegter S, Perna A, Postma MJ, Navis G, Remuzzi G, Ruggenenti P. Sodium intake, ACE inhibition, and progression to ESRD. J Am Soc Nephrol 2012; 23: 165–73.

29 Kang M, Kang E, Ryu H, et al. Measured sodium excretion is associated with CKD progression: results from the KNOW-CKD study. Nephrol Dial Transplant 2021; 36: 512–9.

30 McQuarrie EP, Traynor JP, Taylor AH, Freel EM, Fox JG, Jardine AG, Mark PB. Association between urinary sodium, creatinine, albumin, and long-term survival in chronic kidney disease. Hypertension 2014; 64: 111–7.

31 Fan L, Tighiouart H, Levey AS, Beck GJ, Sarnak MJ. Urinary sodium excretion and kidney failure in nondiabetic chronic kidney disease. Kidney Int 2014; 86: 582–8.

32 Mazarova A, Molnar AO, Akbari A, et al. The association of urinary sodium excretion and the need for renal replacement therapy in advanced chronic kidney disease: a cohort study. BMC Nephrol 2016; 17: 123.

33 Smyth A, O’Donnell MJ, Yusuf S, et al. Sodium intake and renal outcomes: a systematic review. Am J Hypertens 2014; 27: 1277–84.

34 Kieneker LM, Bakker SJ, de Boer RA, Navis GJ, Gansevoort RT, Joosten MM. Low potassium excretion but not high sodium excretion is associated with increased risk of developing chronic kidney disease. Kidney Int 2016; 90: 888–96.

35 Yoon CY, Noh J, Lee J, et al. High and low sodium intakes are associated with incident chronic kidney disease in patients with normal renal function and hypertension. Kidney Int 2018; 93: 921–31.

36 Yuzbashian E, Asghari G, Mirmiran P, Amouzegar-Bahambari P, Azizi F. Adherence to low-sodium Dietary Approaches to Stop Hypertension-style diet may decrease the risk of incident chronic kidney disease among high-risk patients: a secondary prevention in prospective cohort study. Nephrol Dial Transplant 2018; 33: 1159–68.

37 Sugiura T, Takase H, Ohte N, Dohi Y. Dietary Salt Intake is a Significant Determinant of Impaired Kidney Function in the General Population. Kidney Blood Press Res 2018; 43: 1245–54.

38 Ohta Y, Tsuchihashi T, Kiyohara K, Oniki H. High salt intake promotes a decline in renal function in hypertensive patients: a 10-year observational study. Hypertens Res 2013; 36: 172–6.

39 Cirillo M, Bilancio G, Cavallo P, Palladino R, Terradura-Vagnarelli O, Laurenzi M. Sodium intake and kidney function in the general population: an observational, population-based study. Clin Kidney J 2021; 14: 647–55.

40 Institute of Medicine. 2013. Sodium Intake in Populations: Assessment of Evidence. Washington, DC: The National Academies Press.https://doi.org/10.17226/18311.

41 Aburto NJ, Ziolkovska A, Hooper L, Elliott P, Cappuccio FP, Meerpohl JJ. Effect of lower sodium intake on health: systematic review and meta-analyses. BMJ 2013; 346: f1326.

42 Davies NM, Holmes MV, Davey Smith G. Reading Mendelian randomisation studies: a guide, glossary, and checklist for clinicians. BMJ 2018; 362: k601.

43 Munafo MR, Davey Smith G. Robust research needs many lines of evidence. Nature 2018; 553: 399–401.

44 Zhang YM, Zheng J, Gaunt TR, Zhang H. Mendelian Randomization Analysis Reveals a Causal Effect of Urinary Sodium/Urinary Creatinine Ratio on Kidney Function in Europeans. Front Bioeng Biotechnol 2020; 8: 662.

45 Lawlor DA, Tilling K, Davey Smith G. Triangulation in aetiological epidemiology. Int J Epidemiol 2016; 45: 1866–86.

46 Graudal NA, Hubeck-Graudal T, Jurgens G. Effects of low sodium diet versus high sodium diet on blood pressure, renin, aldosterone, catecholamines, cholesterol, and triglyceride. Cochrane Database Syst Rev 2020; 12: CD004022.

47 Huang Y, Wongamorntham S, Kasting J, et al. Renin increases mesangial cell transforming growth factor-beta1 and matrix proteins through receptor-mediated, angiotensin II-independent mechanisms. Kidney Int 2006; 69: 105–13.

48 Ichihara A, Yatabe MS. The (pro)renin receptor in health and disease. Nat Rev Nephrol 2019; 15: 693–712.

49 Aldigier JC, Kanjanbuch T, Ma LJ, Brown NJ, Fogo AB. Regression of existing glomerulosclerosis by inhibition of aldosterone. J Am Soc Nephrol 2005; 16: 3306–14.

50 Olde Engberink RHG, van den Hoek TC, van Noordenne ND, van den Born BH, Peters-Sengers H, Vogt L. Use of a Single Baseline Versus Multiyear 24-Hour Urine Collection for Estimation of Long-Term Sodium Intake and Associated Cardiovascular and Renal Risk. Circulation 2017; 136: 917–26.

51 Dougher CE, Rifkin DE, Anderson CA, Smits G, Persky MS, Block GA, Ix JH. Spot urine sodium measurements do not accurately estimate dietary sodium intake in chronic kidney disease. Am J Clin Nutr 2016; 104: 298–305.

52 Messerli FH, Hofstetter L, Syrogiannouli L, Rexhaj E, Siontis GCM, Seiler C, Bangalore S. Sodium intake, life expectancy, and all-cause mortality. Eur Heart J 2021; 42: 2103–12.

