## SUPPLEMENTAL TABLE OF CONTENTS for "Urinary sodium excretion is not associated with the incidence of end-stage kidney disease and kidney-related death: results from the UK Biobank"

**Figure S1**. Histograms of 24-h urinary potassium excretion calculated by the INTERSALT (a) and Kawasaki (b) equations.

Vertical dashed lines indicate the upper limit of daily sodium intake (2 g/d) recommended by the World Health Organization.

**Table S1**. Incidence rates and adjusted hazard ratios for primary and secondary outcomes according to 24-h urinary sodium excretion with subdistribution hazards models.

**Table S2**. Adjusted hazard ratios for primary outcomes according to 24-h urinary sodium excretion under 3 sensitivity analyses.

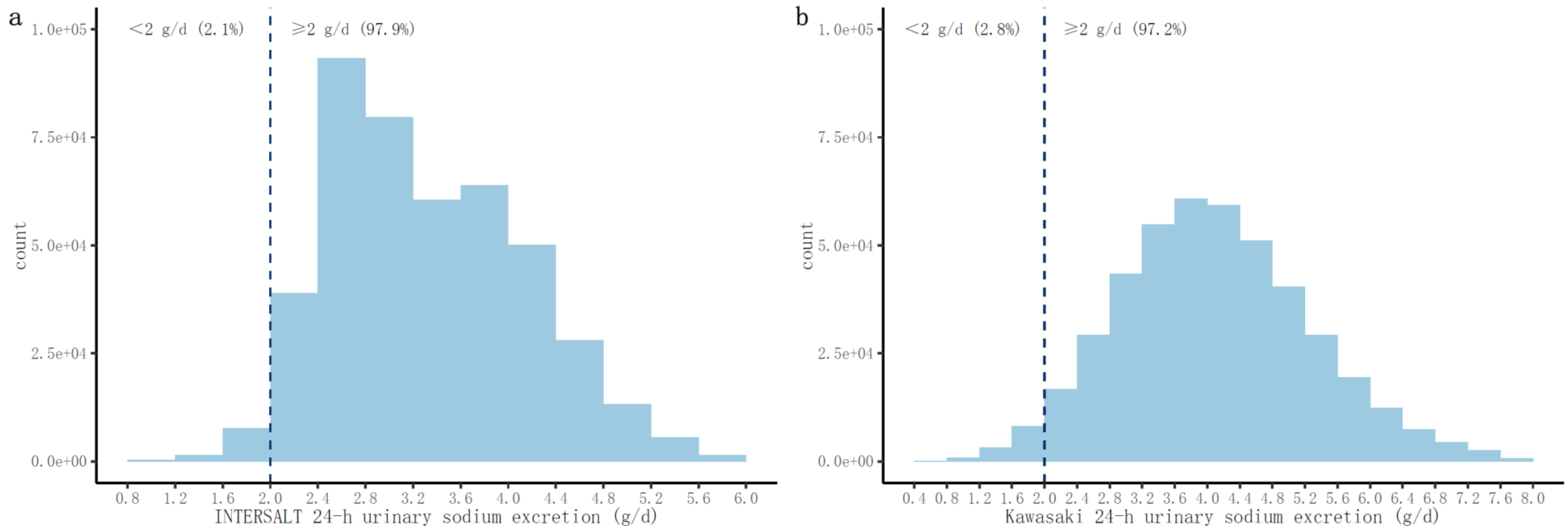


**Figure S1**. Histograms of 24-h urinary potassium excretion calculated by the INTERSALT (a) and Kawasaki (b) equations. Vertical dashed lines indicate the upper limit of daily sodium intake (2 g/d) recommended by the World Health Organization.

**Table S1**. Incidence rates and adjusted hazard ratios for primary and secondary outcomes according to 24-h urinary sodium excretion with subdistribution hazards models.

| | Crude | | Model 1 | | Model 2 | | Model 3 | |
|---|---|---|---|---|---|---|---|---|
| | HRs (95% CIs) | P | HRs (95% CIs) | P | HRs (95% CIs) | P | HRs (95% CIs) | P |
| ***Composite of end-stage kidney disease and kidney-related death*** | | | | | | | | |
| Continuous, per 1 g/d increment | 1.67 (1.55, 1.79) | <0.001 | 1.43 (1.26, 1.62) | <0.001 | 1.03 (0.91, 1.16) | 0.66 | 0.96 (0.84, 1.10) | 0.60 |
| Binary | | | | | | | | |
| Below 2 g/d | Reference | | Reference | | Reference | | Reference | |
| Above 2 g/d | 0.78 (0.55, 1.10) | 0.15 | 0.79 (0.53, 1.18) | 0.25 | 0.84 (0.56, 1.27) | 0.40 | 0.76 (0.49, 1.18) | 0.23 |
| Multicategory | | | | | | | | |
| Quartile 1 | Reference | | Reference | | Reference | | Reference | |
| Quartile 2 | 0.97 (0.80, 1.18) | 0.79 | 1.31 (1.02, 1.66) | 0.03 | 1.03 (0.80, 1.31) | 0.84 | 1.08 (0.83, 1.42) | 0.56 |
| Quartile 3 | 1.51 (1.27, 1.80) | <0.001 | 1.38 (1.01, 1.88) | 0.04 | 1.04 (0.80, 1.36) | 0.76 | 1.06 (0.79, 1.43) | 0.71 |
| Quartile 4 | 2.60 (2.22, 3.06) | <0.001 | 1.87 (1.36, 2.58) | <0.001 | 1.09 (0.82, 1.46) | 0.56 | 1.01 (0.72, 1.41) | 0.96 |
| P for linear trend | <0.001 | | <0.001 | | 0.58 | | >0.99 | |
| P for quadratic trend | <0.001 | | 0.79 | | 0.89 | | 0.42 | |
| ***End-stage kidney disease*** | | | | | | | | |
| Continuous, per 1 g/d increment | 1.57 (1.40, 1.75) | <0.001 | 1.31 (1.08, 1.58) | 0.006 | 1.07 (0.88, 1.30) | 0.48 | 0.93 (0.75, 1.16) | 0.54 |
| Binary | | | | | | | | |
| Below 2 g/d | Reference | | Reference | | Reference | | Reference | |
| Above 2 g/d | 1.63 (0.73, 3.65) | 0.23 | 1.83 (0.67, 4.98) | 0.24 | 2.06 (0.74, 5.75) | 0.17 | 1.68 (0.60, 4.71) | 0.32 |
| Multicategory | | | | | | | | |
| Quartile 1 | Reference | | Reference | | Reference | | Reference | |
| Quartile 2 | 0.92 (0.67, 1.27) | 0.63 | 1.12 (0.77, 1.64) | 0.55 | 0.92 (0.62, 1.38) | 0.69 | 1.02 (0.67, 1.55) | 0.93 |
| Quartile 3 | 1.63 (1.23, 2.16) | <0.001 | 1.25 (0.82, 1.89) | 0.30 | 1.10 (0.73, 1.66) | 0.63 | 1.03 (0.65, 1.62) | 0.91 |
| Quartile 4 | 2.25 (1.74, 2.94) | <0.001 | 1.38 (0.90, 2.12) | 0.14 | 1.08 (0.68, 1.72) | 0.73 | 0.88 (0.53, 1.47) | 0.63 |

| | | | | | | | | |
|---|---|---|---|---|---|---|---|---|
| P for linear trend | <0.001 | | 0.15 | | 0.59 | | 0.68 | |
| P for quadratic trend | 0.04 | | 0.95 | | 0.80 | | 0.50 | |
| ***Kidney-related death*** | | | | | | | | |
| Continuous, per 1 g/d increment | 1.74 (1.60, 1.88) | <0.001 | 1.52 (1.31, 1.76) | <0.001 | 1.00 (0.87, 1.15) | >0.99 | 0.96 (0.82, 1.12) | 0.57 |
| Binary | | | | | | | | |
| Below 2 g/d | Reference | | Reference | | Reference | | Reference | |
| Above 2 g/d | 0.67 (0.47, 0.97) | 0.03 | 0.67 (0.44, 1.02) | 0.06 | 0.67 (0.44, 1.04) | 0.08 | 0.63 (0.39, 1.00) | 0.05 |
| Multicategory | | | | | | | | |
| Quartile 1 | Reference | | Reference | | Reference | | Reference | |
| Quartile 2 | 0.97 (0.77, 1.22) | 0.80 | 1.37 (1.03, 1.83) | 0.03 | 1.03 (0.77, 1.38) | 0.84 | 1.12 (0.82, 1.55) | 0.47 |
| Quartile 3 | 1.55 (1.26, 1.90) | <0.001 | 1.57 (1.06, 2.32) | 0.02 | 1.09 (0.79, 1.50) | 0.59 | 1.17 (0.81, 1.68) | 0.40 |
| Quartile 4 | 2.79 (2.32, 3.36) | <0.001 | 2.19 (1.46, 3.29) | <0.001 | 1.07 (0.75, 1.52) | 0.71 | 1.05 (0.71, 1.57) | 0.79 |
| P for linear trend | <0.001 | | <0.001 | | 0.67 | | 0.78 | |
| P for quadratic trend | <0.001 | | 0.90 | | 0.78 | | 0.25 | |

Model 1 was adjusted for age, sex, Townsend deprivation index, education, ethnicity, smoking status, alcohol consumption, metabolic equivalents, and 24h urinary potassium excretion.

Model 2 was adjusted for age, sex, Townsend deprivation index, education, ethnicity, smoking status, alcohol consumption, metabolic equivalents, 24-h urinary potassium excretion, waist circumference, hypertension, diabetes, cardiovascular disease, stroke, diuretics, angiotensin-converting enzyme inhibitors or angiotensin II receptor blockers, and estimated glomerular filtration rate.

Model 3 was adjusted for age, sex, Townsend deprivation index, education, ethnicity, smoking status, alcohol consumption, metabolic equivalents, 24-h urinary potassium excretion, waist circumference, hypertension, diabetes, cardiovascular disease, stroke, diuretics, angiotensin-converting enzyme inhibitors or angiotensin II receptor blockers, estimated glomerular filtration rate, systolic blood pressure, diastolic blood pressure, and urine albumin-to-creatinine ratio.

Abbreviations: CIs, confidence intervals; HRs, hazard ratios.

**Table S2**. Adjusted hazard ratios for primary outcomes according to 24-h urinary sodium excretion under 3 sensitivity analyses.

| | Sensitivity analysis 1 | | Sensitivity analysis 2 | | Sensitivity analysis 3 | |
|---|---|---|---|---|---|---|
| Scenarios | Excluded participants who took diuretics, angiotensin-converting enzyme inhibitors or angiotensin II receptor blockers, or glucocorticoids at baseline | | Excluded participants who had prevalent congestive heart failure or CKD stage 4-5 at baseline, or developed ESKD or died within 2 years of follow-up | | Used the exposure of 24-h urinary sodium excretion calculated by the Kawasaki equations | |
| Participants included, n | 360,549 | | 440,003 | | 444,328 | |
| | HRs (95% CIs) | P | HRs (95% CIs) | P | HRs (95% CIs) | P |
| Continuous, per 1 g/d increment | 0.92 (0.75, 1.14) | 0.44 | 0.97 (0.84, 1.11) | 0.62 | 1.03 (0.98, 1.10) | 0.25 |
| Binary | | | | | | |
| Below 2 g/d | Reference | | Reference | | Reference | |
| Above 2 g/d | 0.71 (0.40, 1.25) | 0.23 | 0.72 (0.47, 1.10) | 0.13 | 1.22 (0.89, 1.67) | 0.23 |
| Multicategory | | | | | | |
| Quartile 1 | Reference | | Reference | | Reference | |
| Quartile 2 | 0.90 (0.60, 1.36) | 0.62 | 0.99 (0.74, 1.32) | 0.93 | 0.89 (0.73, 1.09) | 0.25 |
| Quartile 3 | 1.03 (0.65, 1.61) | 0.91 | 0.97 (0.71, 1.33) | 0.87 | 1.01 (0.83, 1.23) | 0.93 |
| Quartile 4 | 0.85 (0.51, 1.42) | 0.54 | 0.97 (0.69, 1.38) | 0.88 | 1.08 (0.88, 1.31) | 0.47 |
| P for linear trend | 0.69 | | 0.88 | | 0.29 | |
| P for quadratic trend | 0.75 | | 0.94 | | 0.20 | |

Models for sensitivity analysis 1 were adjusted for age, sex, Townsend deprivation index, education, ethnicity, smoking status, alcohol consumption, metabolic equivalents, 24-h urinary potassium excretion, waist circumference, hypertension, diabetes, cardiovascular disease, stroke, and estimated glomerular filtration rate.

Models for sensitivity analysis 2 and 3 were adjusted for age, sex, Townsend deprivation index, education, ethnicity, smoking status, alcohol consumption, metabolic equivalents, 24-h urinary potassium excretion, waist circumference, hypertension, diabetes, cardiovascular disease, stroke, diuretics, angiotensin-converting enzyme inhibitors or angiotensin II receptor blockers, and estimated glomerular filtration rate.

Note that more participants were included in sensitivity analysis 3 than in the main analysis due to fewer participants with incomplete 24-h urinary sodium excretion data calculated by the Kawasaki equations.
Abbreviations: CIs, confidence intervals; CKD, chronic kidney disease; ESKD, end-stage kidney disease; HRs, hazard ratios.
